# Clinico epidemiological profile of Omicron variant of SARS CoV2 in Rajasthan

**DOI:** 10.1101/2022.02.11.22270698

**Authors:** Ravi Prakash Sharma, Swati Gautam, Pratibha Sharma, Ruchi Singh, Himanshu Sharma, Dinesh Parsoya, Farah Deba, Neha Bhomia, Nita pal, Varsha A Potdar, Pragya D Yadav, Nivedita Gupta, Sudhir Bhandari, Bharti Malhotra

**Author notes:** **Corresponding author and contact information:** Dr. Bharti Malhotra Senior Professor & Head, Microbiology SMS Medical College Jaipur 302004 **email:**, Mob No. **equal contribution from first four authors** (Ravi Prakash Sharma, Swati Gautam, Pratibha Sharma, Ruchi Singh).

## Abstract

**Background:** Omicron a new variant of SARS COV2 was first detected in November 2021. This was believed to be highly transmissible and evade immunity as a result urgent need was felt to screen all positive, identify Omicron cases and isolate them to prevent spread of infection and study their clinico-epidemiological profile.

**Methodology:** All positive cases detected in state of Rajasthan during November to January beginning were selected for next generation sequencing. Processing was done as per protocol on Ion Torrent S5 system for 1210 samples and bioinformatics analysis was done.

**Results:** Among the 1210 samples tested 762(62.9%) were Delta/Delta like and other lineages, 291(24%) were Omicron and 157(12.9%) were invalid or repeat samples. Within a month the proportion of Delta and other variants was reversed, from zero omicron became 81% and delta and other variants 19%, initially all omicron cases were international travelers and their contacts but soon community transmission was seen. Majority of omicron patients were asymptomatic (56.7%) or had mild disease (33%), 9.2% had moderate symptoms and 2(0.7%) had severe disease requiring hospitalization, of which one (0.3%) died and rest (99.7%) recovered. History of vaccination was seen in 81.1%, of previous infection in 43.2%. Among the Omicron cases BA.1 (62.8%) was the predominant lineage followed by BA.2(23.7%) and B.1.529 (13.4%), however rising trends were seen initially for BA.1 and later for BA.2 also.

**Conclusion:** In very short time Omicron has spread in community and has taken over the preexisting Delta/Delta like and other lineages, it evades immunity, but the good part is most of the cases were asymptomatic or had mild disease and mortality rate was very low.

## Background

On November 24^th^, 2021, B.1.1.529 a new variant of SARS CoV2 was reported to the World Health Organization (WHO). This variant was first detected in Botswana on 11^th^ November 2021 and on 14^th^ November 2021 in South Africa which was later termed Omicron by WHO and declared as a Variant of Concern (VOC)eventually (1). Omicron (21 M) or the Pango lineage B.1.1.529 includes 21K omicron (BA.1) and its sister clade 21L Omicron (BA.2) and other diverse Omicron sequences. 21L and 21K share 38 mutations but 21L has additional 27 mutations (with 12 unique mutations) and 21K has 20 more (6 unique deletion/ mutation), 21L lacks the SH69-and Sv70-deletions which lead to S gene drop out or SGTF which has been used a proxy marker for Omicron in Taq Path PCR kits (1).

It’s reported that Omicron spreads 70 times faster than the Delta variant and can evade the immunity provided by natural infection and vaccination due to the mutations which are known to increase transmission, immune escape and enhance binding affinity (2). Though the preliminary data suggest that the illness caused may be asymptomatic to mild disease. But the severity of disease due to Omicron variant remains questionable as many factors like immune status, age comorbid conditions etc may affect it and may vary in different regions. Omicron has been reported from various countries and now from many states of India too. It’s predicted that very soon it will take over the delta strain and become the dominant strain. It’s important to study the clinico-epidemiological profile of the new variant for effective control and treatment. We scaled up the genomic surveillance to include all positives from whole state of Rajasthan and specially the foreign travellers and their contacts at Sawai Man Singh Medical College (SMSMC) Jaipur as satellite lab of the Indian SARS-CoV-2 Genomics Consortium (INSACOG)and National Institute of Virology (NIV)Pune being the hub lab for SMSMC.

### Objective

of the study was to carry out the genomic surveillance of SARS COV2 and study the Clinico-epidemiological profile in patients with Omicron variant.

## Methodology

Nasopharyngeal/Throat swab specimens collected in Viral Transport medium (VTM) from all positive patients of SARS CoV2 at Rajasthan received at SMS Medical College Jaipur from 24^th^ November 2021 to 4^th^ January 2022 for Next Generation Sequencing (NGS) were included in this study. RNA extraction was done on automated extraction system, Nucliesens EasyMAg (BioMérieux, France) using 400ul VTM. Patients were confirmed to be positive for SARS-CoV-2 using TRU PCR (3B Black Bio Biotech, India) real time reverse transcriptase PCR (RT-PCR) kit. Samples,1210 having high viral load (cycle threshold <25 for E and ORF gene) were selected for NGS analysis. Briefly quantification of extracted RNA was done using Qubit HS RNA kit (Life Technologies, USA). Superscript VILO reverse transcriptase kit (Invitrogen, USA) was used for cDNA synthesis. Library preparation was done using Ion AmpliSeq library kit plus (Life Technologies) and Ion AmpliSeq SARS-CoV-2 research assay panel (Life Technologies) as per manufacturer’s instructions. Template preparation and enrichment was done using Ion OT2 kit on Ion OneTouch 2 and Ion OneTouch ES systems (Life Technologies) and then loaded onto an Ion 530 chip for sequencing on the Ion torrent S5 system. The FASTA files obtained on sequencing were downloaded from the Torrent Suite software version 5.0.5 and the reads obtained were then aligned withWuhan-Hu-1/2019 (genbank: MN908947) as reference and the sequences downloaded from Global Initiative on Sharing All Influenza Data (GISAID)using Nextclade (https://clades.Nextstrain.org). Nextclade carries out different processes such as sequence alignment (pairwise alignment using a variation of Smith Waterman algorithm), clade assignment, and phylogenetic placement of the sequences, which can be visualised and or downloaded. The phylogenetic tree was downloaded and visualised using Nextstrain Auspice (accessed on 05^th^ February 2022) web software.

## Results

### Clusters

The first nine Omicron positive cases were seen in a family with history of travel to south Africa and their close contacts. The family of four international travellers reported negative at South Africa and at Dubai before arriving in India, on reaching Rajasthan they visited their relatives. One of the local relative developed mild symptoms and underwent SARS COV2 testing and was found positive which led to testing of the family, another four came positive (total 5) and on tracing their contact history, the international travellers were traced and tested positive (4). These international travellers had gone to a wedding and another of their four contacts who attended the wedding were then found positive. Eventually other members of local family, their driver with his family also became positive for Omicron. Thus affecting 19 persons in cluster earlier samples were of B.1.1.529 lineage and later cases were found to be BA.1. Another big cluster was reported from district jail of nine patients all were found to be BA.2 positive and had mild symptoms.

### History of contact or travel

Among the 291 omicron cases 45 (15.4%) had history of international travel, 33 (11.3%) had history of national travel, 68(23.4%) were the contacts of known positive case. No history of contact or travel was obtained in 145 (49.8%) patients (Table-1). Among international travellers’ maximum were from United Arab Emirates (UAE-24.4%) (Table-2)

**Table 1.** Details of travel & vaccination history in Omicron cases at Rajasthan.

| District | NO | Status of vaccination |  |  |  | Travel History |  |  |  |
| --- | --- | --- | --- | --- | --- | --- | --- | --- | --- |
|  |  | Partially | Fully | Not Eligible | Unvaccinated | International | National | Contact History | Others |
|  | No | No | No | No | No | No | No | No | No |
| Jaipur | 206 | 10 | 173 | 15 | 8 | 23 | 24 | 42 | 117 |
|  | % | **4.8** | **83.9** | **7.3** | **3.8** | **11.1** | **11.6** | **20.3** | **56.8** |
| Ajmer | 20 | 0 | 16 | 4 | 0 | 9 | 2 | 6 | 3 |
|  | % | **0.0** | **80.0** | **20.0** | **0.0** | **45.0** | **10.0** | **30.0** | **15.0** |
| Udaipur | 9 | 0 | 9 | 0 | 0 | 2 | 0 | 4 | 3 |
|  | % | **0.0** | **100** | **0.0** | **0.0** | **22.2** | **0.0** | **44.4** | **33.3** |
| Sikar | 6 | 1 | 3 | 1 | 1 | 0 | 0 | 5 | 1 |
|  | % | **16.6** | **50.0** | **16.6** | **16.6** | **0.0** | **0.0** | **83.3** | **16.6** |
| Bhilwara | 7 | 1 | 3 | 2 | 1 | 2 | 1 | 0 | 4 |
|  | % | **14.3** | **42.8** | **28.5** | **14.3** | **28.5** | **14.3** | **0.0** | **57.1** |
| Jodhpur | 3 | 0 | 3 | 0 | 0 | 2 | 0 | 1 | 0 |
|  | % | **0.0** | **100.0** | **0.0** | **0.0** | **66.6** | **0.0** | **33.3** | **0.0** |
| Alwar | 8 | 1 | 6 | 0 | 1 | 1 | 1 | 1 | 5 |
|  | % | **12.5** | **75.0** | **0.0** | **12.5** | **12.5** | **12.5** | **12.5** | **62.5** |
| Bikaner | 3 | 0 | 2 | 1 | 0 | 0 | 0 | 0 | 3 |
|  | % | **0.0** | **66.6** | **33.3** | **0.0** | **0.0** | **0.0** | **0.0** | **100.0** |
| Pratapgarh | 7 | 0 | 6 | 1 | 0 | 3 | 0 | 4 | 00.0 |
|  | % | **0.0** | **85.7** | **14.3** | **0.0** | **42.8** | **0.0** | **57.1** | **0** |
| Sirohi | 3 | 1 | 1 | 1 | 0 | 3 | 0 | 0 | 0 |
|  | % | **33.3** | **33.3** | **33.3** | **0.0** | **100** | **0.0** | **0.0** | **0.0** |
| Bharatpur | 2 | 0 | 1 | 0 | 1 | 0 | 0 | 0 | 2 |
|  | % | **0.0** | **50.0** | **0.0** | **50.0** | **0.0** | **0.0** | **0.0** | **100.0** |
| Hanumangarh | 1 | 0 | 0 | 0 | 1 | 0 | 0 | 0 | 1 |
|  | % | **0.0** | **0.0** | **0.0** | **100** | **0.0** | **0.0** | **0.0** | **100.0** |
| Kota | 7 | 0 | 5 | 0 | 2 | 0 | 0 | 5 | 2 |
|  | % | **0.0** | **71.4** | **0.0** | **28.5** | **0.0** | **0.0** | **71.4** | **28.5** |
| Jhunjhunu | 1 | 0 | 1 | 0 | 0 | 0 | 0 | 0 | 1 |
|  | % | **0.0** | **100.0** | **0.0** | **0.0** | **0.0** | **0.0** | **0.0** | **100.0** |
| Other | 8 | 1 | 7 | 0 | 0 | 0 | 5 | 0 | 3 |
|  | % | **12.5** | **87.5** | **0.0** | **0.0** | **0.0** | **62.5** | **0.0** | **37.5** |
| **Total** | **291** | **15** | **236** | **25** | **15** | **45** | **33** | **68** | **145** |
|  | % | **5.1** | **81.1** | **8.6** | **5.1** | **15.4** | **11.3** | **23.4** | **49.8** |

**Table 2.** International travel history in Omicron cases.

| Country | Cases |  |
| --- | --- | --- |
| South Africa | 5 | 11.1% |
| Ukraine | 1 | 2.2% |
| Ghana | 1 | 2.2% |
| Nigeria | 4 | 8.9% |
| Zambia | 2 | 4.4% |
| USA | 7 | 15.6% |
| Tanzania | 1 | 2.2% |
| UAE | 11 | 24.4% |
| Spain | 1 | 2.2% |
| UK | 4 | 8.9% |
| Switzerland | 1 | 2.2% |
| Congo | 2 | 4.4% |
| London | 1 | 2.2% |
| Italy | 1 | 2.2% |
| Bangkok | 1 | 2.2% |
| New York | 1 | 2.2% |
| France | 1 | 2.2% |
| Total | 45 | 100.0% |

### Genomic analysis

Among 1210 samples tested, 157(12.9%) samples gave invalid results or were repeat samples (were removed from analysis), 1053(87%) gave good quality results, of which 762 (72.3%) belonged to Delta/Delta Like and other lineages {Delta (218; 20.7%), Delta like (538; 51.1%), No VOC (4; 0.3%), Alpha (1; 0.1%), B.1 (1; 0.1%)}, 291(27.6%) were Omicron. Among the Omicron cases, 39 (13.4%) belonged to B.1.1.529 lineage, 183(62.8%) to BA.1, 69 (23.7%) to BA.2. (Table 3, Figure1).

**Figure 1.**
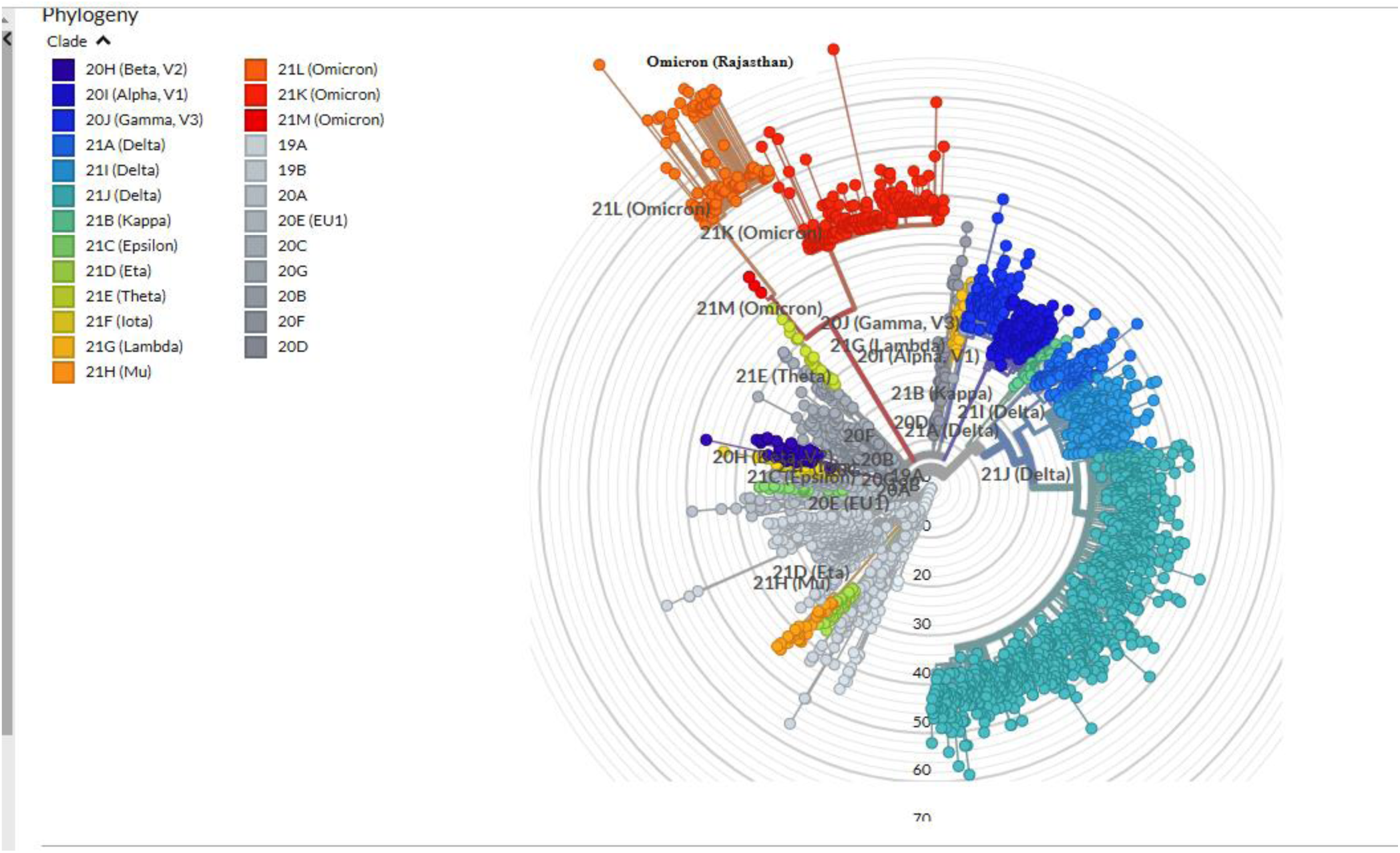
Omicron detected at Rajasthan depicted in Nextclade.

**Table 3.** Week wise distribution of Omicron and other variants in Rajasthan.

| WEEK NO. | valid sequence | Delta, delta like & other variants | totalOMICRON | B.1.1.529 | BA.1 | BA.2 |
| --- | --- | --- | --- | --- | --- | --- |
| 48 | 145 | **136 (93.7)** | **9(6.2)** | **9(100)** |  |  |
| 49 | 239 | 231(96.6) | 8(3.4) | **3(37.5)** | **5(62.5)** |  |
| 50 | 228 | 223(97.8) | 5(2.2) | **4(80)** | **1(20)** |  |
| 51 | 100 | 76(76) | 24(24) | **15(62.5)** | **8(33.3)** | **1(4.1)** |
| 52 | 131 | 56(42.7) | 75(57.2) | **5(6.6)** | **56(74.6)** | **14(18.6)** |
| 1 | 210 | 40(19) | 170 (81) | **3(1.7)** | **113(66.4)** | **54(31.7)** |
|  | 1053 | 762(72.3) | 291(27.6) | 39(13.4) | 183(62.8) | 69(23.7) |

### Week wise positivity

Positivity for Omicron and other variants is given in table 3, in very short time Omicron rose from 0% to 81% overtaking Delta and Delta like variants. Initially the B.1.1.529 lineage was 100% but later both BA.1 and BA.2 emerged and BA.1 was the most predominant but BA.2 was found to be increasing each week. Highest positivity (68 %) was seen in 19-to-59-year age group, 17.1% positivity was seen in paediatric age group also (Table -4) higher positivity was seen in males (56.7%) than females (43.2%). Highest positivity for Omicron was found in Jaipur followed by Ajmer and Udaipur (Table-1),

**Table 4.** Age & sex wise distribution of Omicron cases.

| Age group | Male | % Male | Female | % Female | Total | % |
| --- | --- | --- | --- | --- | --- | --- |
| 0-9 | 4 | 1.40% | 2 | 0.7 | 6 | 2 |
| 10-18. | 20 | 6.90% | 24 | 8.2 | 44 | 15.1 |
| 19-29 | 35 | 12.00% | 29 | 9.9 | 64 | 22 |
| 30-39 | 20 | 6.80% | 19 | 6.5 | 39 | 13.4 |
| 40-49 | 37 | 12.70% | 22 | 7.5 | 59 | 20.3 |
| 50-59 | 19 | 6.50% | 17 | 5.8 | 36 | 12.3 |
| 60-69 | 22 | 7.50% | 6 | 2.1 | 28 | 9.6 |
| 70-79 | 8 | 2.70% | 6 | 2.1 | 14 | 4.8 |
| >=80 | 0 | 0.00% | 1 | 0.3 | 1 | 0.3 |
| **Total** | **165** | **56.70%** | **126** | **43.3** | **291** | **100** |

### Clinical profile

Majority, 56.7% of patients were asymptomatic, 33% had mild disease (sore throat and myalgia) 9.6% had moderate disease/symptoms (fever, myalgia, cough, loss of taste and smell etc), 2(0.7%) patients, of more than 60 years old with multiple comorbidities developed respiratory distress and had to be hospitalised and required oxygen, one of them recovered while the other died in 7 days after illness. Facility quarantine was done for Omicron positive international travellers and their Omicron positive contacts while other patients were home isolated. Time to recovery ranged from 0 -15 days, majority of patients recovered in 7 days’ time (Figure-2), only 1(0.3%) patient died while rest 290 (99.7%) recovered.

**Figure 2.**
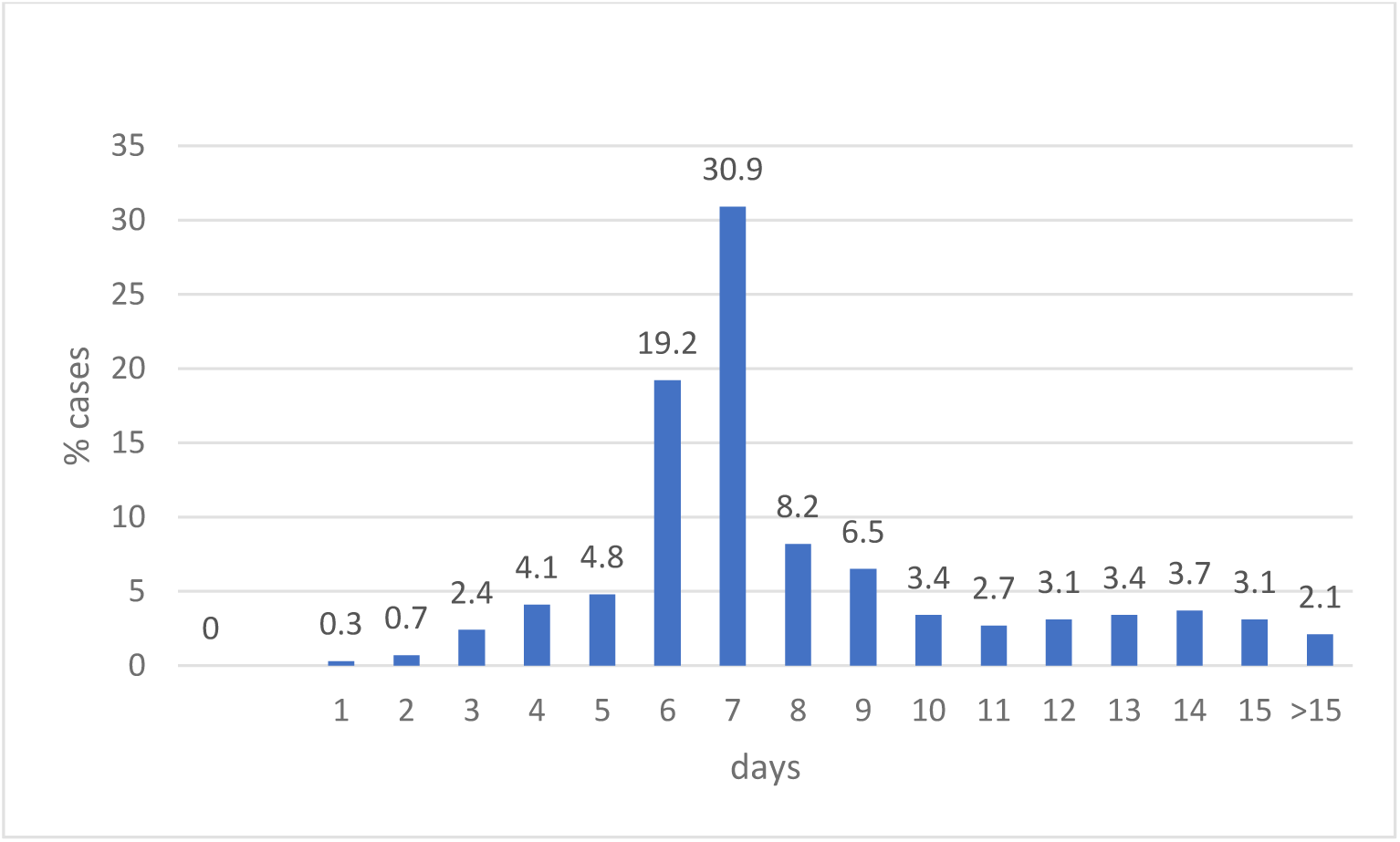
Time to recovery after PCR positivity in Omicron cases.

### Correlation of clinical profile with genomic analysis

On correlation of clinical profile with lineage we found that in asymptomatic patient’s lineage detected was BA.1-31.1%, BA.2-13.4%, B1.1.529-12%, in mild symptom cases BA.1-23%, BA.2-9.2%, B1.1.529-0.7%, in moderate symptoms cases BA.1 was 7.9%, BA.2-1%, B.1.1.529-0.7%, both the two severe disease cases belonged to were BA.1. Since BA.1 was the most predominant lineage it was found to be predominant among all the clinical groups.

### Vaccination status

Details of vaccination status in various districts is given in table 1, 81.1% were fully vaccinated and5.1 % had only one dose, 5.1% were not vaccinated, 8.6% were not eligible for vaccination. Time elapsed between vaccination and RT PCR positivity is given to figure 3, in majority (77.3%) of cases it was less than six months. Majority (70.3 %) of patients had taken Covishield, 20.9 % Covaxin, 6.6% Pfizer and 2.2 % Astra Zeneca and 126 (43.2%) patients had history of past infection in last six months.

**Figure 3.**
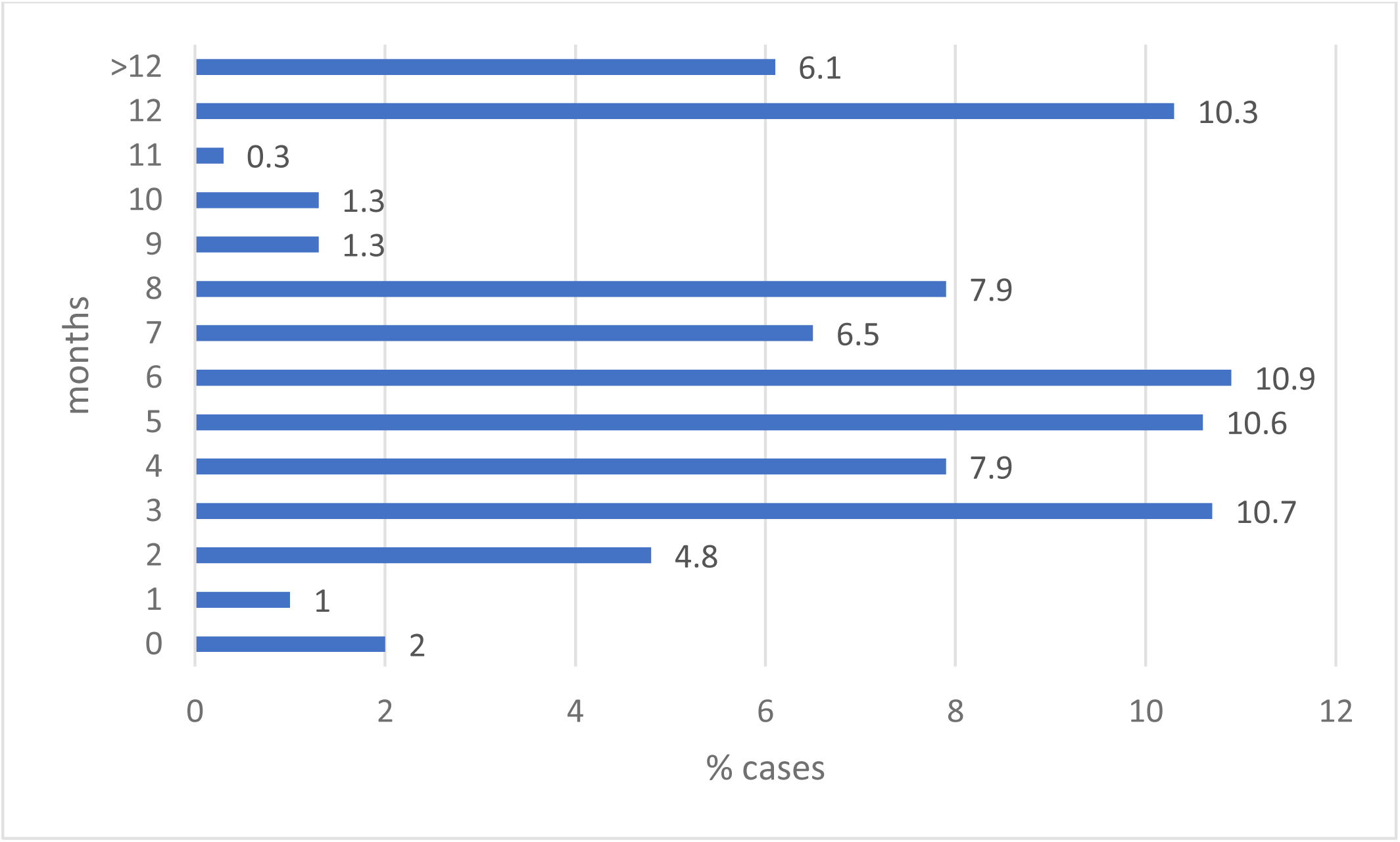
Months elapsed since vaccination & PCR positivity.

### Ethical approval

The study was approved by ethical committee of SMS Medical College Jaipur.

## Discussion

Severe Acute Respiratory Syndrome Coronavirus 2 (SARS-CoV-2) was first isolated from Wuhan, China in December 2019 (4). Soon the virus spread worldwide, and pandemic was declared by World Health Organization in March 2020. Since than due to many mutations various variants have emerged and caused various waves of infection. The second wave in India was caused by Delta variant which caused high morbidity and mortality, increase in number of hospitalisations, high requirement for oxygen and many cases of mucormycosis too. With the emergence of Omicron in South Africa, the concern was its effect on Indian population.

We found first cluster of Omicron in a family of four returning from South Africa and their contacts in 1^st^ week of December which in turn infected 11more cases at Jaipur from their family, their staff, and his family too, later four more from Sikar who had come to attend a wedding in Jaipur, initial cases were of B.1.1.529 lineage while in later contacts BA.1 lineage emerged (5). In our study only 15.4% had history of international and 11.3% of travel in India,23.4% were their contacts, but most importantly 49.8% cases had no history of travel or contact this indicated that community transmission had occurred by late December 2021 to early January 2022.Even another study from Delhi reported community transmission by early January (6). The transmission history stresses the need for social distancing, use of mask and sanitizers and limiting the numbers in public gatherings.

In our study majority of patients were asymptomatic (56.7%) or had mild disease (33%), only 28(9.6%) had moderate disease and only two (0.7%) patients had severe disease requiring hospitalisation. No difference was found in clinical profile of different lineages of Omicron. Majority recovered in seven days’ time except one person who succumbed to death. Though as part of facility quarantine the international travellers and their contacts were initially admitted in designated area of the hospital. As per studies from Gauteng, South Africa only 4.9% case were admitted in hospital in fourth wave which was due to Omicron as compared to 18.9% and 13.7% in third and second waves which were due to Delta and other variants and 28.8% admitted cases in fourth wave had severe disease as compared to 60.1% and 66.9% in third and second waves (7). As per the South African study the proportion of cases admitted in Omicron dominated wave were lower than Delta dominated wave and the sever cases were lower too. However, a study from Imperial college London reports that there is no evidence that the disease severity or hospitalisation due to Omicron are lower than Delta (8). Many factors like age, geographic area, immunisation coverage etc can affect severity of disease. Moreover, there is need for close monitoring of all hospitalised, especially severe cases and deaths during the Omicron driven third wave in India to understand the clinical and public health implication of the new variant.

Omicron is said to be more transmissible than the delta; the scare is that there will be high positivity. The Omicron poses 5.4 times higher risk of reinfection than Delta and the distribution by age, region and ethnicity may also be different in Omicron. Various modelling groups have predicted that SARS CoV2 infections will reach an unprecedented peak in next one to two months and may reach 35 million per day which is triple of the delta wave, its estimated that infection-hospitalisation rate may be 90-96 per cent lower for omicron as compared to delta, and the infection-fatality rate will also be 97-99 per cent lower for the same comparison (9,10,11). Omicron has affected more than 140 countries worldwide and most of the states in India and the numbers of Omicron have also risen to more than 10000. India reported 2.85 lakh new <u>Covid-19</u> cases and 665 deaths in last 24 hours on 26^th^ January 2022 with 22.23 lakh active cases; the daily positivity rate was 16.16% with 93.2% recovery rate. Positivity was seen even when India has already administered 163.58 crore vaccine doses till now. In country like ours if only small percentage gets hospitalised and few die the sheer numbers will be high and will affect the health care systems. Moreover, the need for testing tracing and tracking and providing supportive treatment to mild cases, enhanced infection control measures, quarantine, isolation of infected staff, will greatly affect the health care systems. Since majority of patients were asymptomatic or had mild disease, it will be difficult to trace and track the positive persons as a result it will spread widely in community (2). In our study we found 23.7 % strains to belonged to BA.2, this lineage doesn’t have the SH69- and Sv70- deletions which lead to S gene drop out or SGTF which has been used a proxy marker for Omicron in Taq Path PCR kits therefore using such kits may not detect Omicron moreover we found a rising trend in BA.2 lineage in our samples hence use of these kits can give false negative for Omicron (7,12). However, a new kit Omisure (TATA Medical and Diagnostics) validated by ICMR for Omicron detection may be used to detect omicron which will be particularly useful to identify if the hospitalised patient is omicron or not as with the increase in positivity all the positives cannot be sequenced anymore (13).

In our study 81.1 % of patients were fully vaccinated and 5.1 % were partially vaccinated, time elapsed in 77.3% cases was less than 6 months. Majority (70.3%) of patients had taken Covishield, 20.6% had taken Covaxin while only few had taken other vaccines abroad Pfizer (6.6%), AstraZeneca (2.2%). As per a serosurvey done in Rajasthan during November 2021-December 2021 we observed 85% to 94% population had neutralising antibodies against SARS COV2, even the nonvaccinated had seropositivity of 74% (unpublished data). The low severity observed in our cases may be due to the protective effect of the past vaccination or infection. However, Omicron has been reported to evade the immune response both due to vaccination and earlier infection too. As per a report from Imperial college London the vaccine effectiveness due to Omicron versus Delta variant after two doses of vaccine (AstraZeneca or Pfizer) was 0% and 20% and after booster dose was 55% and 80% respectively (8).As per the vaccine surveillance report from UK after 2 doses of the AstraZeneca vaccine, vaccine effectiveness against the Omicron was initially 45 to 50% but dropped to no effect after 20 weeks of second dose. Similarly, after 2 doses of Moderna or the Pfizer the efficacy reduced from 65% to 10 % by 25 weeks after 2^nd^ dose. However, after the additional booster dose the efficacy dropped from 65% to 25%-40% in 15 weeks, efficacy was better in younger age group than higher age and better in Delta than Omicron (14). An interesting observation in a study done at Pune demonstrated that substantial immune response was seen after omicron breakthrough infection against other variants. The Omicron infected persons sera could neutralise not only the Omicron, but also other variants of concern, including the most prevalent <u>Delta variant</u> thus reducing the chances of reinfection due to Delta and as a result replacing the Delta variant in the population. This stresses the urgent need to have an Omicron-specific vaccine strategy. (15). Though the virus is known to evade the immune response the severity of infection will be lower in the immunised person so it’s important that the booster dose/ precautionary dose is taken timely. Moreover, it’s suspected that non vaccinated may bear the brunt so aggressive drives should be there to vaccinate all.

In nutshell as per our preliminary data the Omicron is highly transmissible; in very short time it has spread in the community and has overtaken the existing Delta strain. It causes mainly asymptomatic to mild disease in vaccinated persons and severe disease in persons with comorbidities. It’s important to plan for Omicron specific vaccination and giving additional booster dose/ precautionary dose to frontline workers and those with comorbidities on priority, carry out sequencing of hospitalised, death cases, nonvaccinated to know the variant responsible among the serious and in unvaccinated cases. It’s important that social distancing and use of mask is ensured for preventing the transmission of infection.

## Data Availability

All data produced in the present work are contained in the manuscript

## Author Contributions

SG, PS, FD, DS, carried out the experiments, SG, PS, BM, VAP, PDY carried out the bioinformatics analysis, RS, RPS, HS carried out data acquisition and compiling of data, NP, NB, HS carried out data compilation, SB, NG carried out funding acquisition and review of manuscript and data, BM carried out conceptualization, manuscript preparation, data analysis, funding acquisition and coordination of the research, RPS, RS, SG, PS, edited the manuscript. All authors have read and agreed to the published version of the manuscript.

## Conflicts of Interest

The authors declare no conflict of interest.

## Ethical approval

The study was approved by the Institutional Ethics Committee of SMS Medical College Jaipur.

## Financial support & sponsorship

Financial support was provided by the Government of Rajasthan and Department of Health Research, Ministry of Health & Family Welfare, New Delhi, to SMS Medical College VRDL Jaipur

## Acknowledgement

We acknowledge the support provided by Shree Vaibhav Galriya Principal Secretary Medical education Rajasthan Government, Dr Balram Bhargav Director General Indian Council of Medical Research, the teams at NIV Pune; NIC team, Bioinformatics team, BSL4 team and Indian SARS-CoV-2 Genomics Consortium (INSACOG).

## Notes

### Competing Interest Statement

The authors have declared no competing interest.

### Funding Statement

This study was funded by state government

### Author Declarations

Ethics committee of SMS Medical College Jaipur approved the study

